# Temporal and frontal lobe contributions to neural synchronization dysfunction and auditory attention in Fragile X Syndrome

**DOI:** 10.1101/2022.05.10.22274929

**Authors:** Ernest V Pedapati, Lauren E. Ethridge, Lisa DeStefano, Yanchen Liu, Makoto Miyakoshi, John A Sweeney, Lauren M. Schmitt, Donald L. Gilbert, Steve W. Wu, Rui Liu, Elizabeth Smith, Rebecca C. Shaffer, Kelli C. Dominick, Paul S. Horn, Devin Binder, Craig A. Erickson

**Author notes:** Corresponding author: Ernest Pedapati, Division of Child and Adolescent Psychiatry, Division of Neurology, Cincinnati Children’s Hospital Medical Center, 3333 Burnet Ave, MLC 4002, Cincinnati, OH 45229. These authors contributed equally to this work (Ernest V Pedapati, Lauren E. Ethridge).

## Abstract

Fragile X syndrome (FXS) is a trinucleotide repeat disorder and the most common hereditary form of intellectual disability. Patients with FXS are commonly impaired by sensory hypersensitivity. Compared to cognition, neural correlates of sensory responses can be studied more objectively and across species. Alterations in auditory processing in FXS measured by scalp electroencephalography (EEG) are well documented. However, the orientation of the auditory cortex in humans introduces significant confounding between temporal and frontal sources. Herein, we conducted EEG source analysis in 36 participants with FXS and 39 matched controls to localize known alterations to the auditory chirp stimulus. The key findings in participants with FXS include 1) distinct contributions of frontal and temporal sources to the onset and intrastimulus neural synchronization, 2) moderation of abnormal neural responses in females with FXS, 3) evidence of deep brain sources contributing to impairments in synchronization, and 4) presence of robust region-specific clinical correlations. The synchronous and asynchronous gamma activity (SA ratio) ratio may further represent a clinically relevant biomarker of the efficiency of auditory processing in FXS. Findings from this study have implications for back translation and relevance for signal-to-noise conceptual frameworks in understanding neural activity in neurodevelopmental disorders.

## INTRODUCTION

Fragile X Syndrome (FXS) is a sex-linked neurodevelopmental disorder caused by a CGG trinucleotide repeat expansion in the *Fmr1* gene, leading to methylation-based silencing of the gene and reduced or silenced translation of fragile X mental retardation protein (FMRP)^1^. Individuals with FXS may show intellectual disability, anxiety, and symptoms of autism, including sensory hypersensitivity and hyperacusis. Sensory hypersensitivity can be particularly distressing for individuals with FXS; however, there are no effective treatments for this aspect of the condition^2^.

Abnormal neural responses to auditory stimuli are robust and reproducible in humans with FXS^3-5^, and in *Fmr1*^-/-^ knock out (KO) mice^6^. Participants with FXS fail to synchronize, measured by intertrial phase coherence (ITPC), to the sensory auditory chirp (0 to 100 Hz linear chirp over 2 s) and exhibit increased asynchronous gamma power^3, 4^. Furthermore, these findings are associated with neurocognitive domains in FXS, including impaired auditory attention, processing speed, and cognitive flexibility^3^. There has been intense interest in investigating the basis of sensory hypersensitivity and hyperacusis. It may reveal fundamental mechanisms of aberrant brain activity in FXS and can be reliability studied across species^2, 7^.

Electroencephalography (EEG) is a highly feasible method of non-invasively probing circuits in clinical populations and can be translated across species. Scalp potentials at each electrode represent the summation of multiple neural sources and, thus, cannot be directly mapped to contributing biological pathways. This is particularly relevant in the auditory domain, where the orientation of the auditory cortex (A1) results in cortical dipoles, which are projected superior and anterior, and thus overlap with tangentially oriented frontal sources. However, this confounding factor can be substantially mitigated using whole-head, dense array EEG. The localization of cortical sources can be estimated by generating a biophysical model. Specifically, by incorporating electrode locations, tissue conductivity, and spatial geometry, a lead-field matrix can be generated to estimate neural activity representing patches of the cerebral cortex^8^.

We undertook source localization of auditory evoked potentials to the chirp stimulus in a cohort of individuals with FXS and matched typically developing controls (TDC) to delineate spatial contribution to auditory processing impairments in FXS. This approach allowed us to investigate cortical substrates associated with early (sensory) versus late (top-down) features, the presence of asymmetry, and contributions of superficial and deep sources related to auditory processing. We hypothesized that neural phase-locking (synchronization to the chirp stimulus) would localize primarily to temporal sources. In contrast, general power differences in low and high frequencies may arise from temporal and frontal sources. These findings have important implications for modeling functional deficits in FXS, providing additional substrate for novel therapeutic development, and enhancing back translation of the development of translational biomarkers.

## MATERIALS AND METHODS

### Participants

An overview of the methods is depicted in **Figure 1**. Thirty-six participants with FXS (23 males; age, 10-53 years; mean age, 25.4±10.3 years) and 39 age- and sex-matched typically developing controls (22 males; age, 12-57 years; mean age, 28.0±12.2 years) participated in this auditory evoked potential (AEP) EEG study (**see Table 1**). Scalp-level electrode analyses of this dataset were published previously^3^. All participants gave written informed consent (or caregiver assent when appropriate), and all procedures were approved by the Cincinnati Children’s Hospital Institutional Board. Mean age and proportion of each sex did not differ between groups, ; howeveras expected, participants with FXS demonstrated a significantly greater burden of clinical symptoms than controls. Control participants were screened for prior diagnosis or treatment for neuropsychiatric illness, medication use, or first-degree relatives affected by serious psychiatric or neurodevelopmental disorders. Participants with FXS were excluded if they had a history of epilepsy or anticonvulsant medications (including benzodiazepines) that could confound EEG interpretation. To ensure a representative sample of the FXS population^9^, participants on stable doses (at least four weeks) of psychiatric medicines were permitted.

**Table 1.** Participant Characteristics: Demographic and clinical features of the EEG dataset, including statistical comparisons between groups. FXS, Fragile X Syndrome; TDC, typically developing control; F, female; M, male; Deviation IQ, Full-Scale IQ, NVIQ, Non-verbal intelligence scale; VIQ, verbal intelligence scale; SCQ, Social Communication Questionnaire; ABC, Aberrant Behavior Checklist; WJ-3, Woodcock III Tests of Cognitive Abilities; ADAMS, Anxiety, Depression, and Mood Scale.

| Measure | Group |  |  |
| --- | --- | --- | --- |
|  | FXS, N = 36<sup>1</sup> | TDC, N = 39<sup>1</sup> | p-value<sup>2</sup> |
| Sex |  |  | 0.5 |
| F | 13 (36%) | 17 (44%) |  |
| M | 23 (64%) | 22 (56%) |  |
| Age at Visit | 25.4±10.3 | 28.0±12.2 | 0.5 |
| Deviation IQ | 42.4±29.1 | 102.9±8.3 | <0.001 |
| NVIQ | 31.9±36.1 | 103.6±11.2 | <0.001 |
| VIQ | 54.9±28.3 | 102.3±10.8 | <0.001 |
| ADAMS General Anxiety | 7.7±4.8 | 2.4±2.6 | <0.001 |
| ADAMS Obsessive/Compliance Behavior | 2.4±2.3 | 0.5±1.3 | <0.001 |
| SCQ Total | 14.0±7.9 | 2.2±2.4 | <0.001 |
| ABC FXS subscale 1: Irritability/aggression | 11.2±11.4 | 0.6±2.4 | <0.001 |
| ABC FXS subscale 4: Hyperactivity/Noncompliance | 7.7±5.9 | 0.9±2.1 | <0.001 |
| ABC FXS subscale 5: Inappropriate speech | 4.4±3.5 | 0.2±0.6 | <0.001 |
| ABC FXS subscale 2: lethargy/social withdrawal | 5.5±4.4 | 0.6±1.5 | <0.001 |
| ABC FXS subscale 3: stereotypy | 4.0±4.5 | 0.1±0.4 | <0.001 |
| WJ-III | 65.2±17.9 | 91.9±11.7 | <0.001 |

<sup>1</sup>n (%); Mean±SD
<sup>2</sup>Pearson's Chi-squared test; Wilcoxon rank-sum test

**Figure 1:**
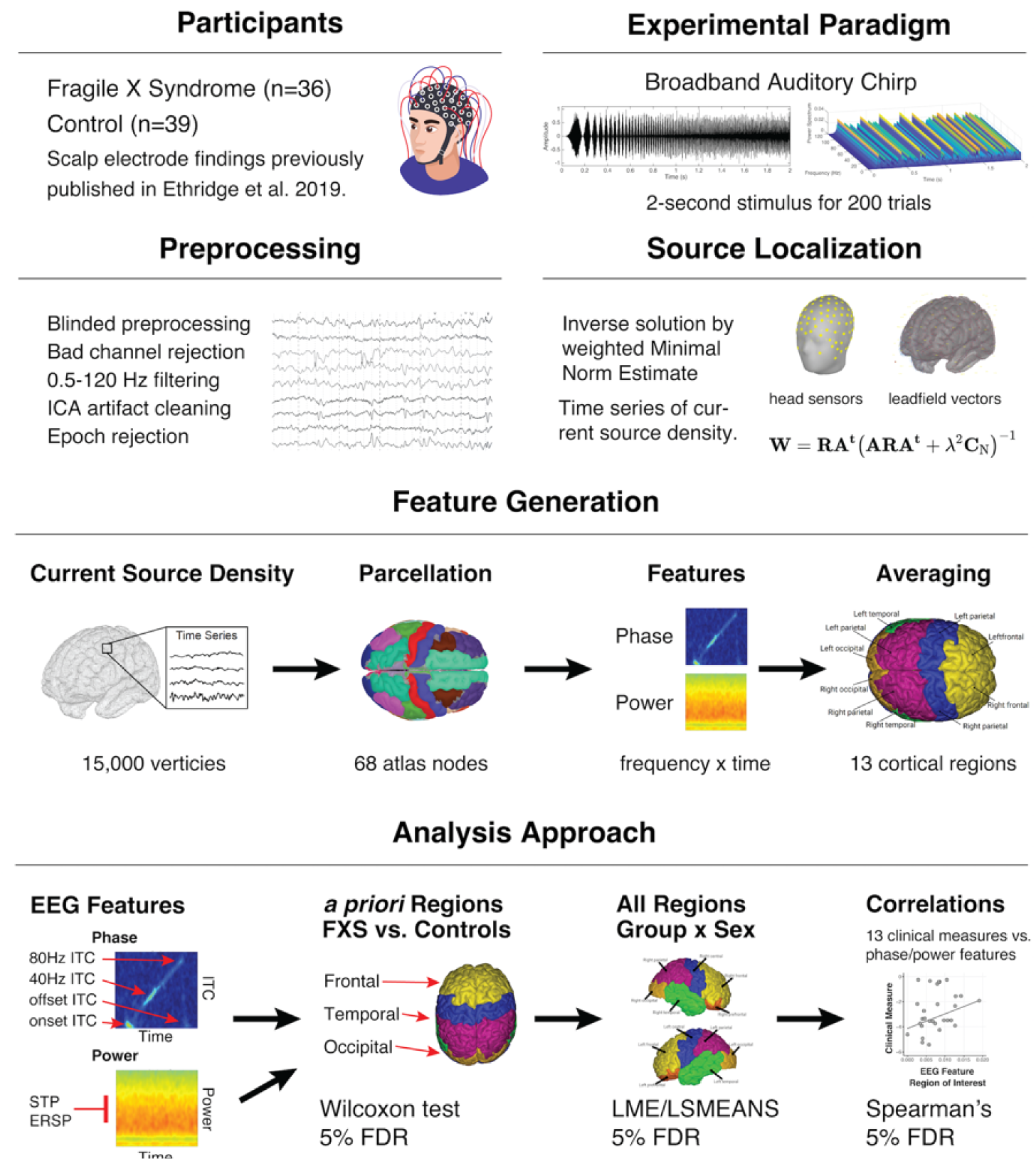
Visual depiction of principal methodology.

### Measures

Caregiver assessments of FXS participants included the Social and Communication Questionnaire (SCQ), Anxiety Depression and Mood Scale^10^ (ADAMS), and Aberrant Behavior Checklist-Community^11^ (ABC-C, optimized for FXS). The participants’ cognitive abilities (IQ) were estimated by the Stanford-Binet Intelligence Scales 5^th^ Ed. Abbreviated IQ. To adjust for floor effects, deviation scores were calculated for verbal and non-verbal IQ in lower ranges^12^.

### Auditory Task

An auditory chirp stimulus can be employed to detect the integrity of functional cortical responses across a wide range of input frequencies (**Supplemental Figure 2A**)^13^. In controls, a strong cortical response to the chirp stimulus is observed and can be quantified by inter-trial phase coherence. The present auditory stimulus consisted of a pink noise carrier amplitude modulated by a sinusoid linear chirp which increased in frequency from 0-100 Hz over 2000 milliseconds (ms) (**Supplemental Figure 2B**). No onset or offset ramps were included. Participants were presented with 300 stimulus repetitions separated by a 1500-2000 ms variable interstimulus interval (randomly jittered).

### Stimulus Presentation

Auditory stimuli were presented through Sony MDR-V150 headphones (65 dB SPL) using Presentation 22 (Neurobehavioral Systems) in a sound-attenuated room. A silent video was used during the task to facilitate protocol compliance and promote quiet wakefulness state^4, 14^.

### EEG acquisition

EEG was continuously recorded referenced to Cz with a sampling rate of 1000 Hz using a NetAmp 400 (Magstim/EGI, Eugene, Oregon) with Hydrocel 128-channel geodesic saline-based electrode net.

### EEG preprocessing

Raw data were visually inspected offline. Data were digitally filtered from .5-120 Hz (12 and 24 dB/octave roll-off, respectively; zero-phase; 60 Hz notch). Bad sensors were interpolated (no more than 5% per subject, no more than two adjacent) using spherical spline interpolation implemented in BESA 6.1 (MEGIS Software, Grafelfing, Germany). Ocular, cardiac, and muscle movement artifacts were removed blind to the participant group using independent components analysis (extended infomax) implemented in EEGLAB (Delorme and Makeig, 2004) using MATLAB 2018a (MathWorks, Natick, MA). Segments of data with large amounts of movement artifact were removed before ICA to facilitate algorithm convergence. Data were then transformed to average reference and epoched into 3250 ms trials (−500 ms to 2750 ms). Any trial with post-ICA amplitude exceeding 120 µV was considered a residual artifact and removed.

### Source analyses

A distributed source model was estimated using a depth-weighted minimum norm estimate (MNE) model in Brainstorm^15^. Various methods have been developed to solve the ill-posed problem of calculating dipolar magnitudes from scalp potentials and electrode positions^16^. An MNE solution identifies a single solution that minimizes total brain power and corresponds to Tikhonov regularization. As the classical MNE solution biases weaker surface sources closer to electrodes, a depth weighting (order, .5; maximal amount, 10) was performed to normalize by a measure of their overall amplitude^17^. To optimize for dense array EEG analysis, the model was constrained to dipoles normal to the cortical surface, and an identity matrix was used for noise covariance. Scalp electrodes were co-registered to an averaged Montreal Neurologic Institute ICBM152 common brain template. The forward model, or lead field matrix, was computed using Open MEEG resulting in a boundary element method head model, which accounts for the brain, CSF, skin, and skull conductivity properties^18^. An MNE kernel was then implemented and used to generate a current source density (CSD) time series across a resultant triangular mesh (15,002 vertices) representing the cortical envelope. The vertices were parcellated into 68 cortical nodes according to the Desikan-Killiany (DK) atlas^19^. The DK atlas also includes thirteen hierarchical anatomical grouping variables into continuous cortical regions (**see Supplemental Figure 2**). Temporal sources include atlas nodes on or adjacent to A1 (banks of superior temporal sulcus, superior temporal gyrus, and transverse temporal gyrus).

### Time-frequency Analyses

For each atlas node, single-trial waveforms were analyzed using the *newtimef* function included with EEGLAB 2021. Convolution parameters included using Morlet wavelets with one Hz frequency steps using a linearly increasing cycle length from one cycle at the lowest frequency (2 Hz) and 30 cycles with the highest frequency (120 Hz). The function calculates three main features: ITPC, single-trial power (STP), and event-related spectral perturbation (ERSP)^20^. ITPC estimates the degree of phase-locked responses across trials; however, it can be biased by trial number. To avoid bias, raw ITPC values were corrected for trial numbers by subtracting the critical r value as follows:

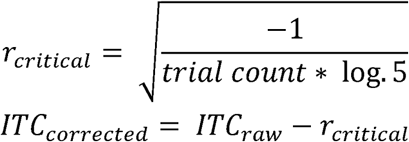

STP measures the spectral power of the frequency response while ERSP quantifies the change in spectral power by frequency corrected for the pre-stimulus baseline (−500 to 0 milliseconds).

### Statistical Analyses

All statistical analysis was performed in R (version 4.1)^21^. Mean STP and ERSP were computed across alpha (8-12 Hz), gamma1 (30-60 Hz), and gamma2 (60-100 Hz) frequency bands. ITPC was examined at several pre-defined time-frequency clusters^3^ corresponding to stimulus onset and offset (alpha frequencies) and 40 Hz and 80 Hz (approximately gamma1 and gamma2 frequency bands). Data analysis was carried out in two stages to confirm previous scalp electrode findings and to investigate new localization-related hypotheses. First, we examined simple group differences in mean response variables within three source regions of interest (ROI): temporal, frontal, and occipital regions (where no task-related activity was expected). Following normality testing (see Supplemental Table 1), we performed Wilcoxon Signed Rank tests comparing FXS to TDC for each ROI for ten response variables. Multiple comparisons were controlled using a false discovery rate (FDR) of 5%.

### Linear modeling

Next, we conducted a series of linear mixed-effects models (NLME library in R 4.1) to examine the effect of group, sex, and region (including right and left regions) on the response variables. After examining various link functions, response variables were log-transformed and raised to 1/3 power. This was validated using Akaike’s information criterion (AIC), examining plots of the distribution of the residuals, residuals over predicted values, and quantile-quantile plots. Fixed effects included GROUP (FXS vs. TDC), SEX (male vs. female), and REGION (13 ROIs). For each REGION, DK atlas nodes were treated as replicates (see Supplemental Table X). Clinical Correlations: Spearman’s rho was used to estimate the association between clinical measures and the EEG response variables. For our primary correlation analysis, we examined relationships across all FXS participants and applied a 5% FDR correction. We additionally examined uncorrected exploratory correlations within a subgroup consisting of males with FXS.

## RESULTS

### Neural response to auditory chirp is significantly altered in FXS

In the temporal region, participants with FXS showed decreased ITPC to the chirp stimulus at 40 Hz, Z=395, FDR-adjusted (adj.) p <.001, and 80 Hz, Z=376, adj. p <.001 (**Figure 2B**). In addition, compared to TDC, FXS gamma1 STP was elevated, Z=1,005, adj. p=.004, and alpha STP was reduced, Z=928, adj. p=.04. In the frontal region, an increase in ITPC onset was observed in the FXS group, Z=990, adj. p=.02. No significant differences in response variables were found in the occipital region. Heat plot visualization of ITPC response is displayed in **Figure 3A-B**. A summary of measures, including effect sizes, is presented in **Table 2**. The results replicate differences in scalp electrode responses to the auditory chirp^3, 4^ but delineate contributions to specific features associated with the frontal and temporal lobes.

**Figure 2:**
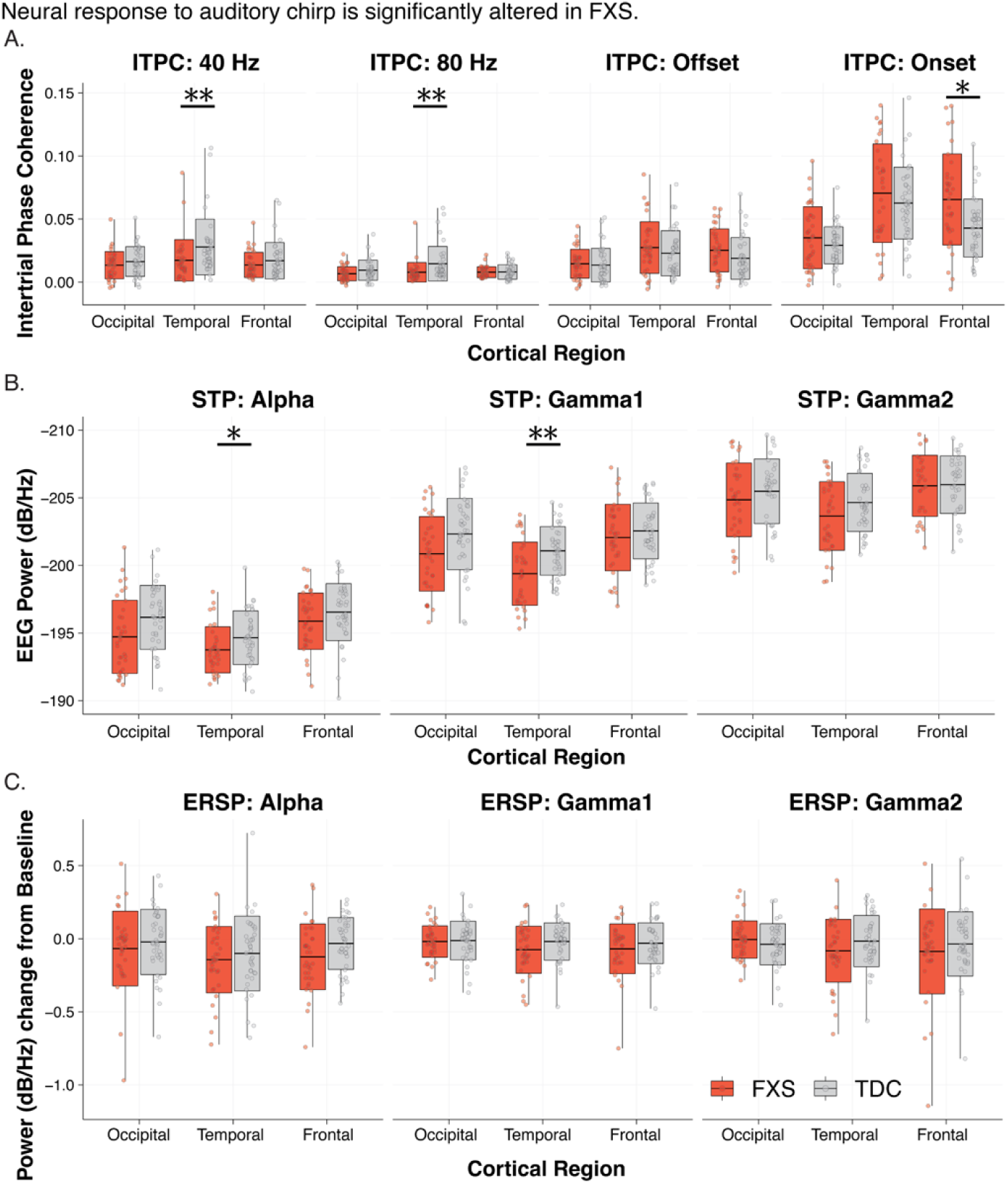
Source-localized auditory evoked potentials to the auditory chirp delineate contributions to auditory processing abnormalities associated with frontal and temporal lobes. A. Intertrial Phase Coherence. B. Single trial spectral power. C. Event-related spectral perturbation.

**Table 2.**
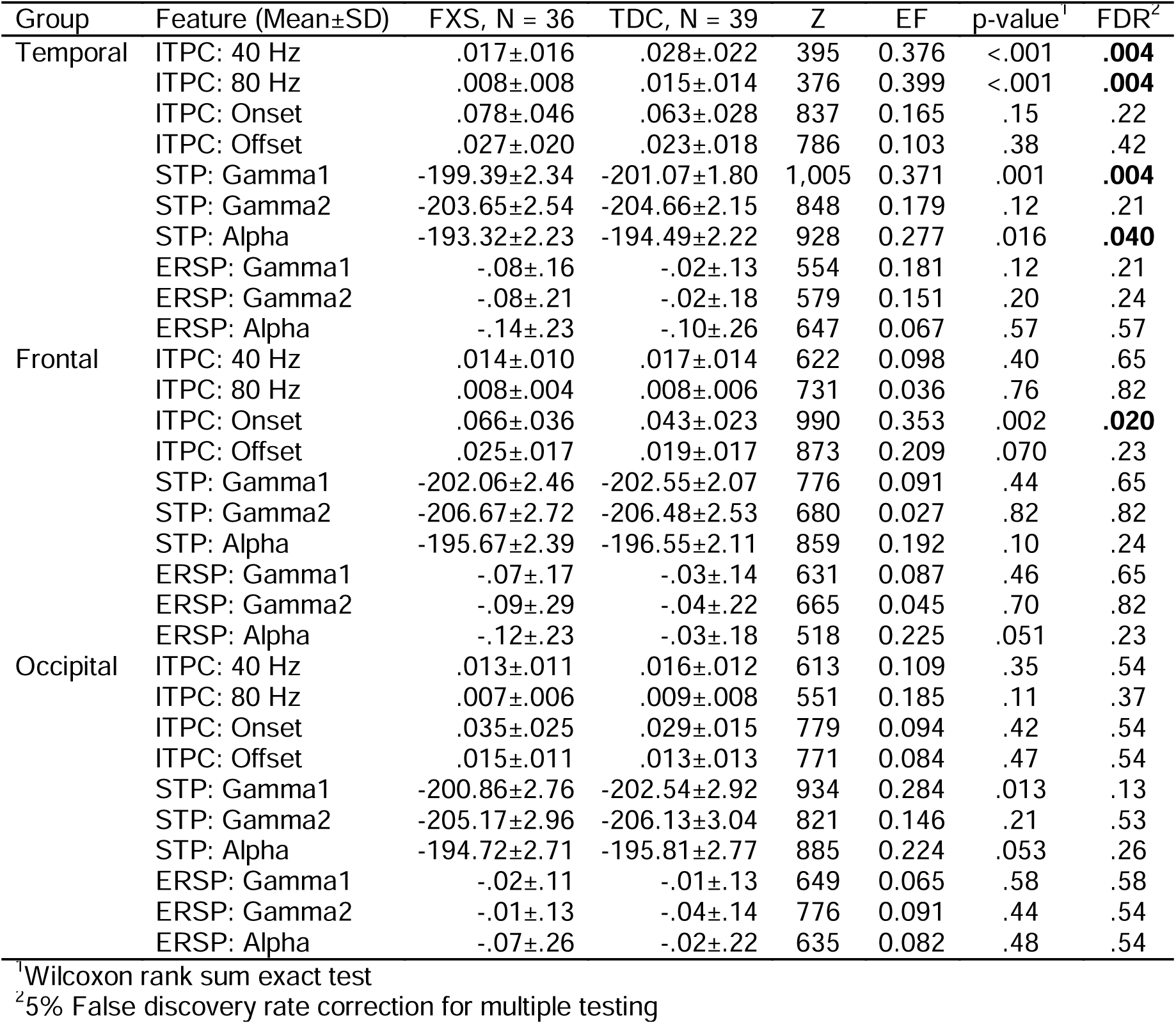
Group differences in Chirp Response across Temporal, Frontal, and Occipital Regions

| Group | Feature (Mean±SD) | FXS, N = 36 | TDC, N = 39 | Z | EF | p-value<sup>1</sup> | FDR<sup>2</sup> |
| --- | --- | --- | --- | --- | --- | --- | --- |
| Temporal | ITPC: 40 Hz | .017±.016 | .028±.022 | 395 | 0.376 | <.001 | **.004** |
|  | ITPC: 80 Hz | .008±.008 | .015±.014 | 376 | 0.399 | <.001 | **.004** |
|  | ITPC: Onset | .078±.046 | .063±.028 | 837 | 0.165 | .15 | .22 |
|  | ITPC: Offset | .027±.020 | .023±.018 | 786 | 0.103 | .38 | .42 |
|  | STP: Gamma1 | -199.39±2.34 | -201.07±1.80 | 1,005 | 0.371 | .001 | **.004** |
|  | STP: Gamma2 | -203.65±2.54 | -204.66±2.15 | 848 | 0.179 | .12 | .21 |
|  | STP: Alpha | -193.32±2.23 | -194.49±2.22 | 928 | 0.277 | .016 | **.040** |
|  | ERSP: Gamma1 | -.08±.16 | -.02±.13 | 554 | 0.181 | .12 | .21 |
|  | ERSP: Gamma2 | -.08±.21 | -.02±.18 | 579 | 0.151 | .20 | .24 |
|  | ERSP: Alpha | -.14±.23 | -.10±.26 | 647 | 0.067 | .57 | .57 |
| Frontal | ITPC: 40 Hz | .014±.010 | .017±.014 | 622 | 0.098 | .40 | .65 |
|  | ITPC: 80 Hz | .008±.004 | .008±.006 | 731 | 0.036 | .76 | .82 |
|  | ITPC: Onset | .066±.036 | .043±.023 | 990 | 0.353 | .002 | **.020** |
|  | ITPC: Offset | .025±.017 | .019±.017 | 873 | 0.209 | .070 | .23 |
|  | STP: Gamma1 | -202.06±2.46 | -202.55±2.07 | 776 | 0.091 | .44 | .65 |
|  | STP: Gamma2 | -206.67±2.72 | -206.48±2.53 | 680 | 0.027 | .82 | .82 |
|  | STP: Alpha | -195.67±2.39 | -196.55±2.11 | 859 | 0.192 | .10 | .24 |
|  | ERSP: Gamma1 | -.07±.17 | -.03±.14 | 631 | 0.087 | .46 | .65 |
|  | ERSP: Gamma2 | -.09±.29 | -.04±.22 | 665 | 0.045 | .70 | .82 |
|  | ERSP: Alpha | -.12±.23 | -.03±.18 | 518 | 0.225 | .051 | .23 |
| Occipital | ITPC: 40 Hz | .013±.011 | .016±.012 | 613 | 0.109 | .35 | .54 |
|  | ITPC: 80 Hz | .007±.006 | .009±.008 | 551 | 0.185 | .11 | .37 |
|  | ITPC: Onset | .035±.025 | .029±.015 | 779 | 0.094 | .42 | .54 |
|  | ITPC: Offset | .015±.011 | .013±.013 | 771 | 0.084 | .47 | .54 |
|  | STP: Gamma1 | -200.86±2.76 | -202.54±2.92 | 934 | 0.284 | .013 | .13 |
|  | STP: Gamma2 | -205.17±2.96 | -206.13±3.04 | 821 | 0.146 | .21 | .53 |
|  | STP: Alpha | -194.72±2.71 | -195.81±2.77 | 885 | 0.224 | .053 | .26 |
|  | ERSP: Gamma1 | -.02±.11 | -.01±.13 | 649 | 0.065 | .58 | .58 |
|  | ERSP: Gamma2 | -.01±.13 | -.04±.14 | 776 | 0.091 | .44 | .54 |
|  | ERSP: Alpha | -.07±.26 | -.02±.22 | 635 | 0.082 | .48 | .54 |

<sup>1</sup>Wilcoxon rank sum exact test
<sup>2</sup>5% False discovery rate correction for multiple testing

**Figure 3:**
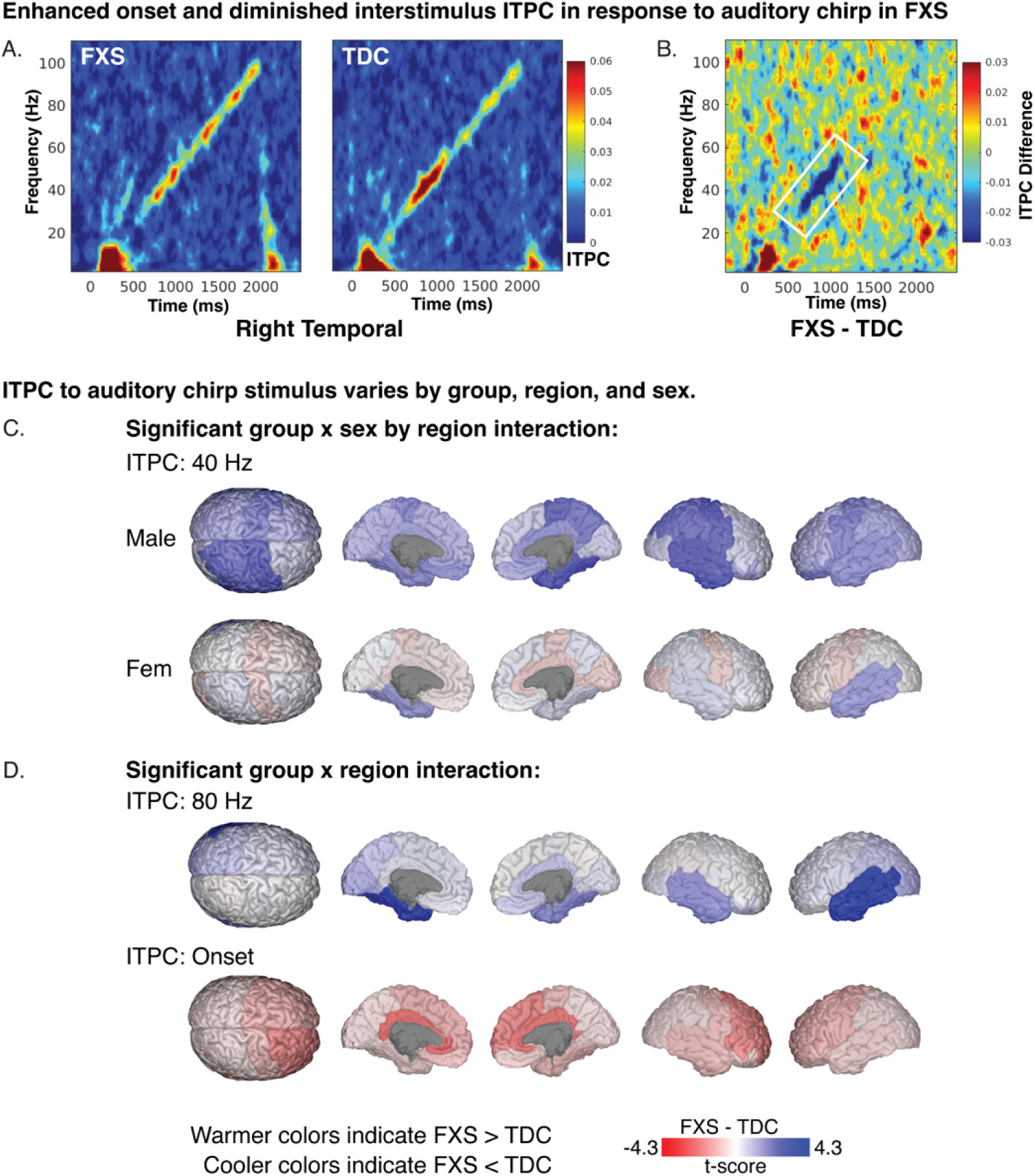
Time frequency heatmaps representing intertrial coherence (ITPC) in response to auditory chirp. A. Plots depicting mean ITPC response by group in the right temporal region. B. ITPC difference plot (FXS-TDC) with white outline marking a significant reduction of 40 Hz ITPC in FXS. C. Brain plots depicting least-square means contrasts (t-values) of 40 Hz ITPC. D. Brain plots depicting least-square means contrasts (t-values) of 80 Hz and onset ITPC.

### Neural response to auditory chirp varies by group, region, and sex

We had specific hypotheses which would be best tested within the source space and summarized in **Table 3**. As females with full mutation FXS are obligate mosaics, studying the effect of sex (in a matched cohort) could reveal physiological differences associated with FXP expression. Furthermore, in studying source localized data, we can examine features of chirp response across cortical areas as well as study the symmetry of the response across hemispheres. We constructed linear mixed effect models to examine the effect of group, sex, and region on the chirp response. Model results are summarized in **Supplemental Table 3**.

**Table 3.**
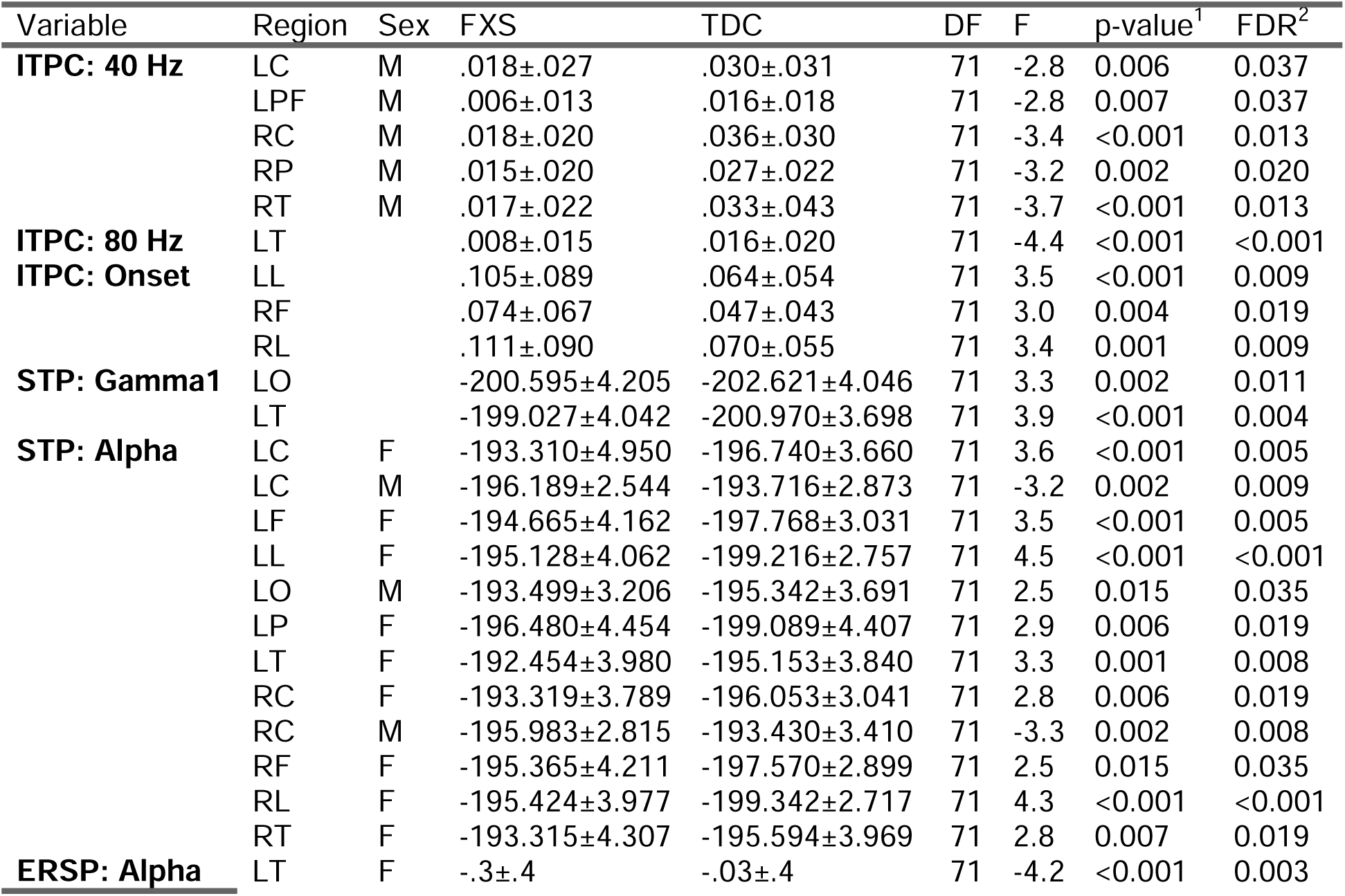
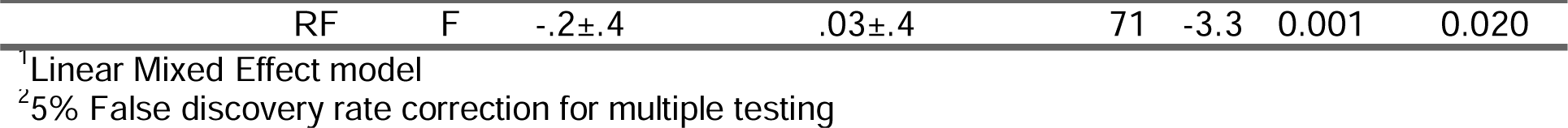
Significant linear mixed model post-hoc contrasts of least-squared means following 5% FDR correction of EEG response variables.

### Enhanced onset and diminished intrastimulus ITPC in FXS

Previous reports have demonstrated a marked reduction of 40 Hz ITPC during chirp stimulus^3, 4^ in human participants as well as local field potentials in *Fmr1*^*-/-*^ KO mice^6^. **Figure 3** summarizes ITPC findings. We found a significant interaction effect of group, sex, and region (F_13,4973_=1.9, p < .001) on 40 Hz ITPC. Following 5% FDR correction, we found group effects were primarily driven by male participants with FXS demonstrating reduced ITPC across the bilateral central region, right parietal, left prefrontal, and right temporal cortex. For 80 Hz ITPC, two-way interaction between group and region was significant (F_13,4973_=4.6, p < .001). Post-hoc comparisons revealed decreased synchronization of FXS (.008±.015) compared to TDC (.016±.020) in the left temporal region (5% FDR p <.001). Two 2-way interactions between group x region (F_13,4973_=2.9, p < .001) and sex x region (F_13,4973_=2.8, p < .001) were significant. Based on our *a priori* hypotheses, we examined post-hoc pairwise contrasts for group x region contrasts. Increased onset ITPC in participants with FXS was found across bilateral limbic and right frontal regions. A significant three-way interaction of group x sex x region was found for offset ITPC, however, no post-hoc pairwise contrasts survived the 5% FDR comparison. A full listing of significant pairwise contrast of ITPC measures is listed in **Table 3**. To better visualize gradients and asymmetry in ITPC response across the cortex, we plotted model-specific post-hoc t-values of region comparisons on a three-dimensional brain atlas. For 40 Hz ITPC, we observed a broad reduction in ITPC in males with FXS compared to control males across the cortex, with the peak difference in the right temporal region. In contrast, females with FXS did not reach statistical significance following a 5% FDR correction in any region (**Figure 3C**). For 80 Hz ITPC, reduction in ITPC was primarily localized to the temporal regions at the group level with the largest difference in the left temporal region (**Figure 3D, Top**). Visualization of the group effect of onset ITPC (FXS > TDC) demonstrated a frontal effect (peak difference in the right frontal region) and limbic regions (**Figure 3D, Bottom**). As the precision of EEG source localization drops precipitously with depth, the limbic regions primarily represent non-superficial sources of deeper brain structures^8, 22^.

### Elevation in asynchronous single-trial gamma power

At the scalp level, we have previously reported increased background gamma power and decreased alpha ERSP in participants with FXS during the chirp stimulus. We wanted to ascertain using source localization associations of spectral power changes with temporal or frontal regions. For alpha STP, a significant group x region x sex interaction effect was present (F_13,4973_=9.6, p <.001). Males with FXS demonstrated a significant decrease in background alpha power across bilateral central regions and the left occipital following correction (**Figure 4D, Top)**. In contrast, females with FXS demonstrated an increase in alpha power across multiple bilateral cortical regions including central, frontal, and limbic regions (**Figure 4D, Bottom**). For gamma1 and gamma2 STP, multiple two-way interactions were significant but based on our hypotheses we examined contrasts for significant group x region interaction effect (gamma1 STP: F_13,4973_=2.6, p =.002; gamma2 STP: F_13,4973_=2.5, p < .002). Following 5% FDR correction, only significant pairwise contrasts were identified for gamma1 STP including elevated gamma power in the left temporal and left occipital regions (**see Figure 4E**).

**Figure 4:**
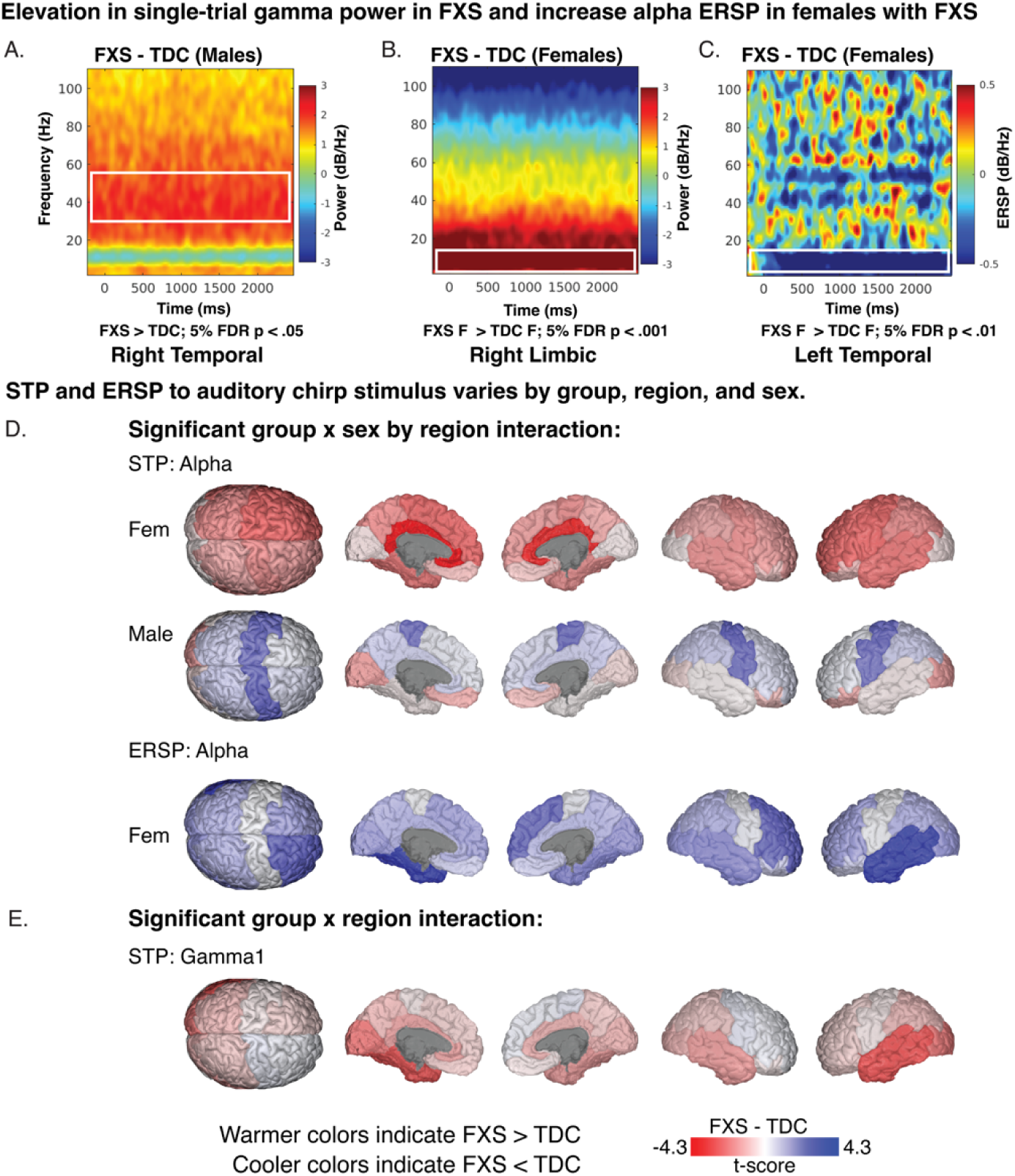
Time-frequency. heatmaps representing single-trial power (STP) and event-related spectral perturbation (ERSP) during auditory chirp. A-C. Exemplar plots visualizing STP and ERSP contrasts (FXS-TDC) which are associated with significant linear mixed model contrasts. D-E. Brain visualizations depicting least-square means contrasts (t-values) of STP and ERSP of significant interaction effects.

**Figure 5:**
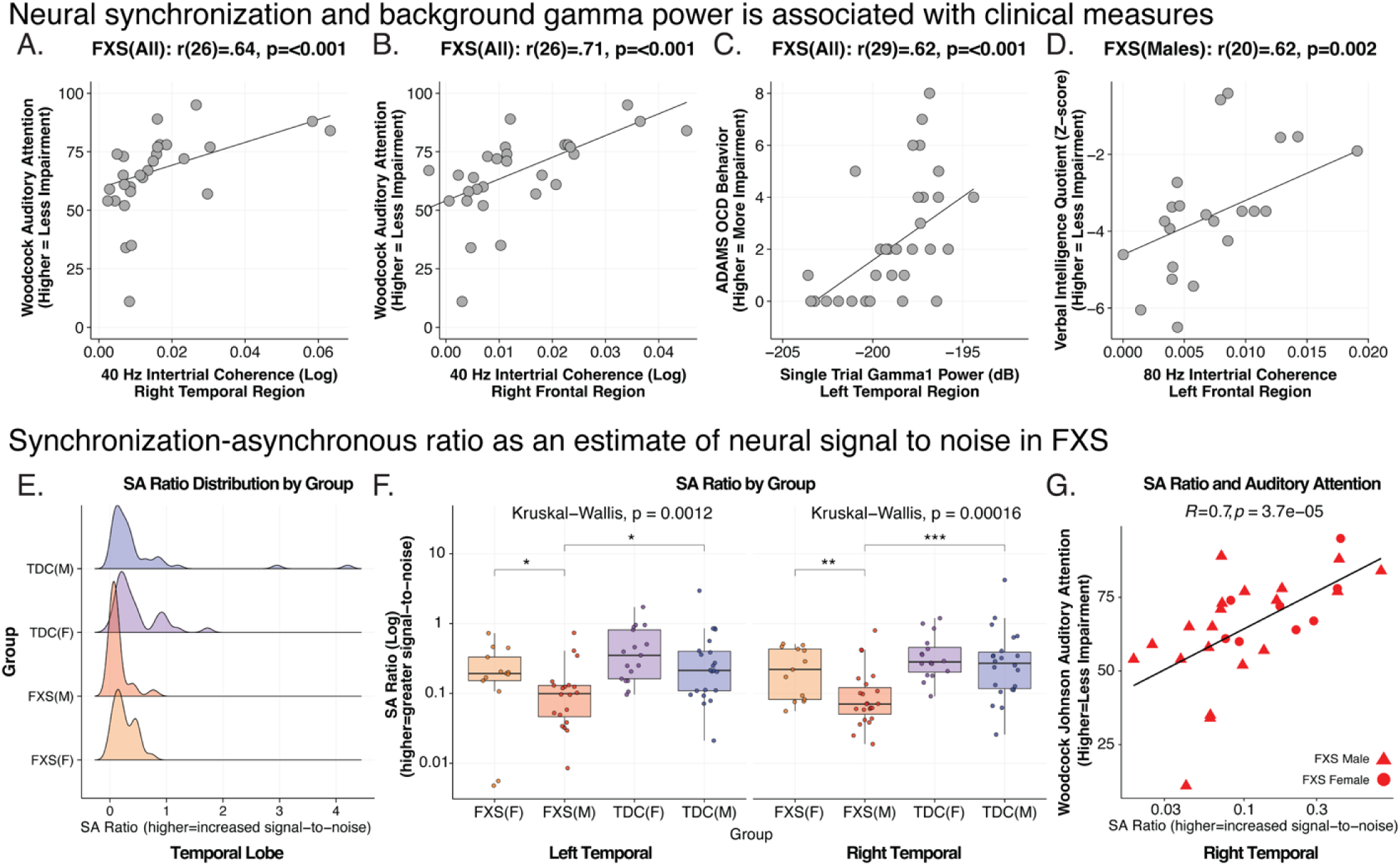
Neural synchronization and background gamma power are associated with clinical measures. A-D. Exemplar clinical correlations with EEG variables across all Fragile X Syndrome (FXS) subjects and a subgroup of full mutation males. Scatterplots in each box depict subject-level bivariate Spearman’s correlations. Synchronization-asynchronization ratio (SA ratio) as an estimate of neural signal to noise (SNR) in FXS. SA ratio is a composite measure of the degree of 40 Hz (gamma) neural synchronization per amount of background gamma noise. E. Distribution of SA in males with FXS is skewed towards lower SA ratio (reduced SNR). F. SA ratio in males with FXS is significantly reduced in bilateral temporal lobes compared to either control males or females with FXS. G. SA ratio is highly correlated with auditory attention performance, such that greater SNR is associated with better auditory attention performance.

### Increased alpha ERSP in females with FXS

Previously, we did not identify significant group differences in ERSP (baseline corrected STP) elicited by the chirp stimulus at the electrode level. A significant group x sex x region interaction was found for gamma1 ERSP (F_13,4973_=2.7, p <.001) and gamma2 ERSP (F_13,4973_=2.0, p =.017). No pairwise contrasts survived 5% FDR correction. For alpha ERSP, a significant group x sex x region interaction was found (F_13,4973_=2.8, p < .001). Corrected pairwise contrasts demonstrated a significant decrease in alpha ERSP following chirp stimulus in females with FXS in the left temporal and right frontal regions. Visualization of t-value region differences between FXS and control females show a broader trend of decreased alpha ERSP across multiple cortical regions (**Figure 4C**).

### Clinical Correlations

We first examined 5% FDR corrected Spearman’s correlations between EEG measures and clinical measures across all FXS participants. We next examined exploratory clinical correlations in males with FXS who are expected to have significantly less FXP protein expression compared to females with FXS^23^. All FXS (see Table X): The WJ3 was used to quantify selective auditory attention (including speech-sound discrimination and resistance to auditory stimulus), with higher scores indicating better performance^24^. Increased performance on the WJ-III was associated with higher 40 (right frontal: r(27)=.71; p<.001, 5% FDR p=.017, right temporal: r(27)=.64; p<.001, 5% FDR p=.035) and 80 Hz ITPC (right temporal: r(27)=.65; p<.001, 5% FDR p=.035). To assess anxiety and obsessive-compulsive symptoms in individuals with intellectual disabilities, we examined the relationship of response variables with the Anxiety, Depression, and Mood Scale (ADAMS) anxiety and obsessive subscales. Higher levels of obsessive-compulsive symptoms were associated with higher gamma1 STP (left temporal: r(30)=.62; p<.001, 5% FDR p=.035). and gamma2 STP (left temporal: r(30)=.64; p<.001, 5% FDR p=.035). The Social Communication Questionnaire is a brief screening instrument to evaluate social-communication skills where higher values indicate greater impairment ^25^. We found trend level associations (10% FDR) such that decreased 40 Hz ITPC (left temporal: r(30)=-.58; p<.001, 5% FDR p=.083) and increased gamma2 STP r(30)=.64; p<.001, 5% FDR p=.083) were associated with higher levels of social communication impairment. Males with FXS (see Table 5): Increased WJ-III performance was associated with increased 40 Hz ITPC across multiple cortical regions with a prominent association with the right frontal region (r(19)=.72; p<.001). In addition, higher left temporal 40 Hz ITPC was associated with higher non-verbal and full-scale (deviation) IQ scores. Increased left frontal 40 and 80 Hz ITPC was correlated with increased verbal IQ. Onset ITPC of bilateral frontal and right temporal regions were associated with ABC measures of inappropriate speech. Higher levels of gamma1 and gamma2 ERSP were associated with worsening ADAMS anxiety scores and OCD scores and ABC inappropriate speech. ERSP alpha power was inversely related with age at visit.

### Synchronization-asynchronous ratio an estimate of neural signal to noise in FXS

We hypothesized the relationship between neural synchronization and background gamma power could be used as a measure of signal-to-noise (SNR). We calculated the ratio of 40 Hz ITPC and gamma1 STP which we termed the synchronous-to-asynchronous ratio (SA ratio). A higher ratio indicates more stimulus phase locking (“signal”) in the gamma frequency range compared to background gamma power (“noise”).

Thus, higher values would indicate a higher proportion of “noise” to “signal” (see Equation 1).

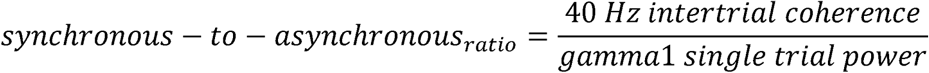

The SA ratio was calculated within each of the thirteen cortical regions for each subject. To examine differences between participant groups (group x sex) we conducted Kruskal-Wallis rank sum tests for each region. The corrected results (5% FDR), as shown in Table 4, indicate SA ratio was significantly different between groups across all regions. To further validate our hypothesis, we predicted that higher SA ratio (i.e., increased SNR) would be associated with improved with auditory attention. As seen in Figure X, we found a significant positive correlation between SA ratio and WJ3 across FXS participants (R=.7, p=3.7e-5).

**Table 4.**
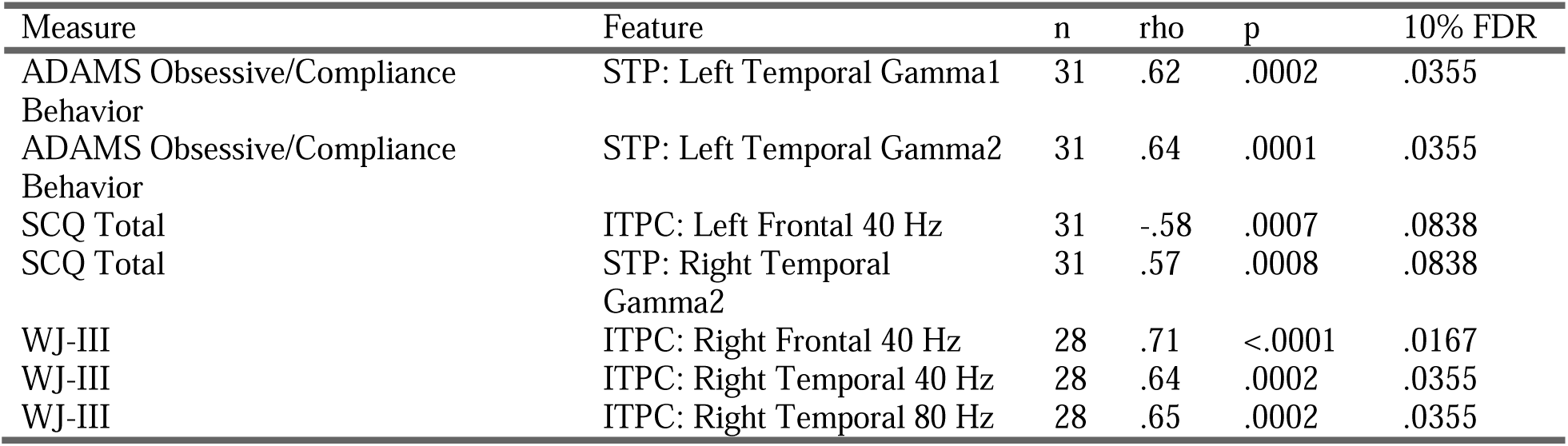
Clinical correlations with Chirp EEG features across all FXS participants.

| Measure | Feature | n | rho | p | 10% FDR |
| --- | --- | --- | --- | --- | --- |
| ADAMS Obsessive/Compliance Behavior | STP: Left Temporal Gamma1 | 31 | .62 | .0002 | .0355 |
| ADAMS Obsessive/Compliance Behavior | STP: Left Temporal Gamma2 | 31 | .64 | .0001 | .0355 |
| SCQ Total | ITPC: Left Frontal 40 Hz | 31 | -.58 | .0007 | .0838 |
| SCQ Total | STP: Right Temporal Gamma2 | 31 | .57 | .0008 | .0838 |
| WJ-III | ITPC: Right Frontal 40 Hz | 28 | .71 | <.0001 | .0167 |
| WJ-III | ITPC: Right Temporal 40 Hz | 28 | .64 | .0002 | .0355 |
| WJ-III | ITPC: Right Temporal 80 Hz | 28 | .65 | .0002 | .0355 |

**Table 4:**
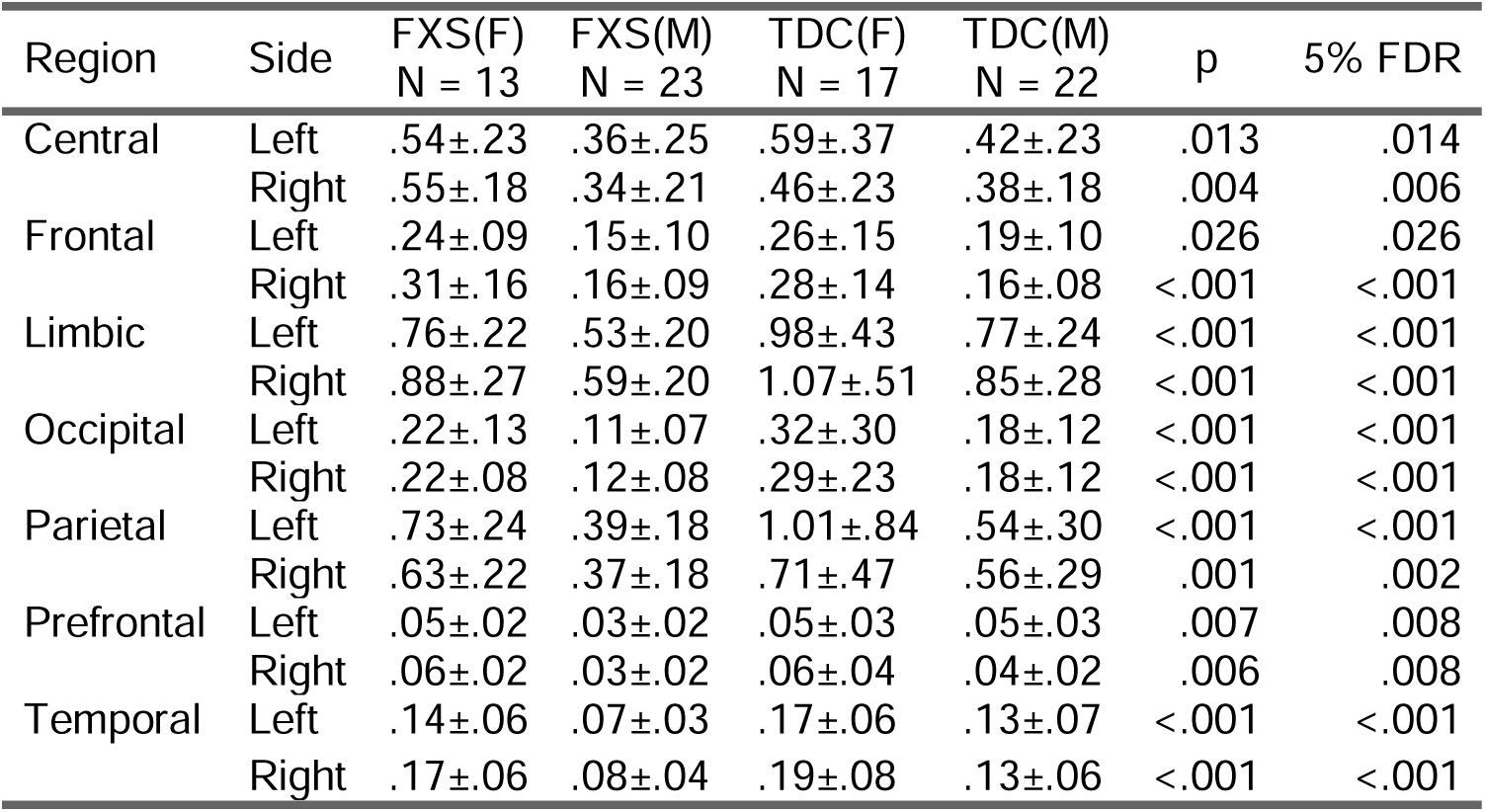
The synchronous-to-asynchronous ratio (SA ratio) was developed as a putative signal-to-noise measure to estimate the efficiency of auditory processing in FXS. Descriptive statistics and corrected mean comparisons of SA ratio are tabulated by cortical region.

| Region | Side | FXS(F)<br>N = 13 | FXS(M)<br>N = 23 | TDC(F)<br>N = 17 | TDC(M)<br>N = 22 | p | 5% FDR |
| --- | --- | --- | --- | --- | --- | --- | --- |
| Central | Left | .54±.23 | .36±.25 | .59±.37 | .42±.23 | .013 | .014 |
|  | Right | .55±.18 | .34±.21 | .46±.23 | .38±.18 | .004 | .006 |
| Frontal | Left | .24±.09 | .15±.10 | .26±.15 | .19±.10 | .026 | .026 |
|  | Right | .31±.16 | .16±.09 | .28±.14 | .16±.08 | <.001 | <.001 |
| Limbic | Left | .76±.22 | .53±.20 | .98±.43 | .77±.24 | <.001 | <.001 |
|  | Right | .88±.27 | .59±.20 | 1.07±.51 | .85±.28 | <.001 | <.001 |
| Occipital | Left | .22±.13 | .11±.07 | .32±.30 | .18±.12 | <.001 | <.001 |
|  | Right | .22±.08 | .12±.08 | .29±.23 | .18±.12 | <.001 | <.001 |
| Parietal | Left | .73±.24 | .39±.18 | 1.01±.84 | .54±.30 | <.001 | <.001 |
|  | Right | .63±.22 | .37±.18 | .71±.47 | .56±.29 | .001 | .002 |
| Prefrontal | Left | .05±.02 | .03±.02 | .05±.03 | .05±.03 | .007 | .008 |
|  | Right | .06±.02 | .03±.02 | .06±.04 | .04±.02 | .006 | .008 |
| Temporal | Left | .14±.06 | .07±.03 | .17±.06 | .13±.07 | <.001 | <.001 |
|  | Right | .17±.06 | .08±.04 | .19±.08 | .13±.06 | <.001 | <.001 |

**Table 5:** Clinical correlations with Chirp EEG features across Male FXS participants.

| Measure | Feature | n | rho | p |
| --- | --- | --- | --- | --- |
| ABC FXS subscale 2: lethargy/social withdrawal | ITPC: Offset(RT) | 20 | -.49 | .0266 |
| ABC FXS subscale 4: Hyperactivity/Noncompliance | ITPC: 80 Hz(LF) | 20 | -.53 | .0174 |
| ABC FXS subscale 5: Inappropriate speech | ERSP: Gamma2(RT) | 20 | -.49 | .0298 |
|  | ITPC: Onset(LF) | 20 | .49 | .0284 |
|  | ITPC: Onset(RF) | 20 | .55 | .0128 |
|  | ITPC: Onset(RT) | 20 | .47 | .0368 |
| ADAMS General Anxiety | ERSP: Gamma1(LO) | 21 | .45 | .0421 |
|  | ITPC: Onset(RT) | 21 | .50 | .0224 |
| ADAMS Obsessive/Compliance Behavior | ERSP: Gamma1(RF) | 21 | -.46 | .0367 |
|  | ITPC: Offset(RT) | 21 | -.51 | .0173 |
| Age at Visit | ERSP: Alpha(LF) | 23 | -.48 | .0198 |
|  | ITPC: 40 Hz(RO) | 23 | .46 | .0263 |
|  | ITPC: 40 Hz(RT) | 23 | .43 | .0412 |
| Deviation IQ | ITPC: 40 Hz(LT) | 22 | .44 | .0423 |
| Non Verbal Z Score | ITPC: 40 Hz(LT) | 22 | .48 | .0244 |
| Verbal Z Score | ITPC: 40 Hz(LF) | 22 | .45 | .0348 |
|  | ITPC: 80 Hz(LF) | 22 | .62 | .0021 |
| WJ-III | ERSP: Gamma2(LT) | 20 | .50 | .0253 |
|  | ERSP: Gamma2(RO) | 20 | -.47 | .0373 |
|  | ITPC: 40 Hz(LO) | 20 | .52 | .0193 |
|  | ITPC: 40 Hz(LT) | 20 | .52 | .0175 |
|  | ITPC: 40 Hz(RF) | 20 | .72 | .0003 |
|  | ITPC: 40 Hz(RO) | 20 | .55 | .0118 |
|  | ITPC: 40 Hz(RT) | 20 | .65 | .0017 |
|  | ITPC: 80 Hz(RT) | 20 | .60 | .0054 |

## DISCUSSION

FXS is primarily marked by cognitive impairments, however, studying sensory responses can provide insight into aberrant brain mechanisms and is readily investigated across species. To date, abnormalities in the neural processing of auditory stimuli using scalp EEG are well documented in FXS and are associated with clinical symptomatology^3-5, 26, 27^. However, due to the orientation of Heschl’s gyrus in humans, electrical potentials relevant to auditory processing are primarily projected through frontal regions, thus posing a confound to mapping results to a cortical substrate. Herein, we performed source modeling to study neural phase and power responses to the auditory chirp. The chirp stimulus can rapidly probe oscillatory response to a broad range of presentation frequencies and results in characteristic bands of phase alignment (or ITPC) at 40 and 80 Hz. The key findings in participants with FXS include 1) distinct contributions of frontal and temporal sources to alterations in neural synchronization, 2) moderation of abnormal neural responses in females with FXS, 3) evidence of deep brain sources contributing to impairments in synchronization, and 4) presence of robust regional clinical correlations.

### Reconciling Spatial Ambiguity by Source Localization

Disentangling temporal and frontal signals has several ramifications for understanding and advancing knowledge of cortical physiology in FXS. We hypothesized that since ITPC is closely linked to basic sensory function, ITPC differences would be most prominent in temporal cortex near A1, whereas absolute power differences may be more diffuse. This hypothesis was partially supported, in that significant group differences in 40 and 80 Hz ITPC were localized to the temporal regions. However, the exaggerated onset ITPC appeared to be primarily a frontal phenomenon. To date, it is not well understood to what extent known frontal abnormalities in FXS^28-33^ impact auditory processing. Functional and structural changes suggesting delayed frontal lobe maturation have been reported in FXS^29, 34, 35^ and frontal pathways contributing to auditory responses have been described^36-38^. In the present study, we found that deficits in neural synchronization (40 Hz and 80 Hz ITPC) were predominately localized to the temporal regions, whereas significant increases in onset ITPC were localized to the frontal and limbic (deeper) regions.

### Reduced Neural Synchronization Localized to Temporal Regions in FXS

As ITPC is most closely linked to basic sensory function, we confirmed our hypothesis that deficits in neural synchronization in FXS were localized to temporal regions, close to A1. The reduction in 40 Hz ITPC was primarily driven by males with FXS in the right temporal region. In contrast, both males and females with FXS had reduced 80 Hz ITPC compared to TDC in the left temporal. We had a relatively strict correction of 5% FDR to establish significant findings, but trend-level differences (10% FDR) were in line with our hypothesis. For example, 40 Hz and 80 Hz ITPC demonstrated trend-level differences in FXS from TDC in the contralateral temporal regions. The auditory pathway from brainstem to the cortex is particularly rich in FXP-expressing neurons^39^ and FXP loss alters the properties of auditory neurons^40, 41^. Thus, loss of FXP is predicted to have a large and sex-dependent impact on auditory processing and may underlie why auditory hypersensitivity is a common phenotype in humans.

### Translational parallels of chirp response in FXS

Several aspects of the chirp response in humans are recapitulated in the Fmr1^-/-^ KO. Forty and 80 Hz ITPC are significant reduced in Fmr1^-/-^ KO mice with differences found across frontal, medial, and temporal regions^6^. Why is the effect more widespread in the mouse than humans? It is expected that the human sample is genetically more heterogeneous, including mosaic males and females (who are obligate mosaics) and increased FXP production should lead to normalization of brain signals. In addition, non-trivial amounts of peripheral FXP have been observed in a proportion of non-mosaic full mutation males^23^. Second, ignoring nomenclature, scalp potentials in mice, as in humans, represent a mixture of source activity at each electrode. Interestingly, murine EEG synchronization to chirp stimulus remains intact following deletion of *Fmr1* from cortical excitatory neurons. This suggests that FXS loss from other cell types or subcortical circuits are necessary to observed EEG abnormalities. The use of depth electrodes in the mouse model, which is not feasible at scale in humans, may provide a more definitive method of source localization. The combination of surface and depth electrodes may lead to a better understanding of cortical (and possibly subcortical) drivers of alterations in auditory processing.

### Exaggerated Stimulus Onset Localized to Frontal Regions in FXS

Unexpectedly, we found the exaggerated onset ITPC to the chirp^3, 4^ was localized to the right frontal region in FXS. As participants watched a silent movie to facilitate compliance with the task and divert attention from the stimulus, we had expected reduced influence of frontal top-down modulation. In males with FXS, this increased onset response was associated with inappropriate speech and anxiety. Furthermore, in males with FXS 40 Hz and 80 Hz synchronization deficits in the frontal regions were associated with worse verbal intelligence, auditory attention, and hyperactivity/impulsivity. It is known, for example, those temporal lobe regions associated with early and late auditory processing project to frontal regions^38^. Exaggerated frontal activation on stimulus onset in FXS may generally represent increased excitability or reduced inhibition of frontal circuits such that early sensory afferent signals are not effectively filtered. Methodology between species will need to more closely aligned to draw further mechanistic insight into the observed frontal response – to date, murine studies to date have employed a lead-in ramp to the chirp stimulus which attenuates the onset response.

Nevertheless, sensory responses in humans are likely a more complex phenomena and there is a need to clarify mechanistic contrasts with animal models. For example, as *Fmr1*^*-/-*^ KO mice also demonstrate reduced ERP habituation to repeated tones^6^ and increased spiking of auditory cortical neurons to sound^42^. However, mice also display audiogenic seizures, which there is no direct homolog in humans (AGS)^43^. In mice, brainstem alterations have been observed in excitatory neurons which are necessary and sufficient to replicate the AGS phenotype. However, contrary to expectation, no changes have been observed in auditory brainstem responses in human patients with FXS^44, 45^. Thus, we raise speculation that auditory hypersensitivity in FXS, which also involve psychological and behavioral manifestations in humans, may be the results of temporal and frontal contributions.

### Rigid gamma circuits in FXS

How can we reconcile increased ITPC during stimulus onset, but reduced ITPC at 40 Hz and 80 Hz frequencies observed in FXS? Findings from the *Fmr1*^*-/-*^ KO suggest stimulus-independent hypersynchrony in the gamma frequency range may be inflexible to phase resetting by incoming stimuli. Recordings within layer 4 of the *Fmr1*^*-/-*^ KO somatosensory cortex demonstrate disordered firing patterns and reduced gamma synchrony to thalamic stimulation^46-48^. Interestingly, stimulus-independent hyper-synchrony of gamma oscillations has been observed between layers 2/3 and 5 of the *Fmr1*^*-/-*^ KO auditory cortex^49^. These findings are consistent with excessive gamma activity recorded at the scalp in *Fmr1*^*-/-*^ KO^6^ and individuals with FXS^14, 50^, including the present study. Stimulus-locked gamma oscillations play an important role in filtering and amplifying of sensory stimuli^51, 52^. Accordingly, if there is abnormal inflexibility in gamma modulation in FXS, we would predict an excessive response to sensory onset, but also diminished 40 Hz and 80 Hz neural synchronization, which is what we have observed.

Elevations of broadband gamma activity in FXS has been well-documented at rest and during tasks^14, 50, 53^ and may represent a dynamic phenomenon. In the present data, we isolated the most significant elevations in background gamma (i.e., STP) to the left temporal regions in FXS. Though other areas (such as the right temporal) reached trend level significance, gamma activity was not universality elevated across sources. Taken as a whole, the data suggests that gamma elevation in FXS is not fixed to a particular region but may be associated to task activity. Furthermore, we have recently shown, that more complex auditory tasks elicit group differences in gamma power in frontal sources^28^. We also recently identified increased gamma activity in the visual cortex of participants with FXS during quiet wakefulness and viewing a silent video. Thus, though basal gamma activity in FXS may be modestly altered, neural processing of cognitive and sensory demands may be more significantly affected.

### Contribution of deep brain sources to neural synchronization

Females with FXS demonstrated distinct abnormalities in the alpha band during stimulus presentation. Background alpha power was markedly elevated in deeper sources compared to controls; however, alpha power was also more strongly desynchronized in females with FXS. Though this finding was unexpected, as females are obligate mosaics, the results can provide insight into FXS physiology. Typically, alpha oscillations are the dominant rhythm at rest and desynchronize following auditory stimulation^54, 55^. However, when a stimulus is presented against background noise, alpha power appears prominently increased in occipital and temporal regions^56, 57^. This effect appears to be independent of ITPC and presumed to be a form of selective inhibition of task irrelevant sensory information^58, 59^. Alpha activity can be viewed as a continuum, such that low alpha power is associated with continuous cortical processing, but high alpha power states can lead to inhibition of cortical circuits though alpha phase dynamics^60^. Recently, we detected a marked reduction in resting absolute and relative alpha power in males with FXS, but relatively spared in females^50, 61^. Our conclusion suggested that males with FXS have evidence of thalamocortical dysfunction which leads to slower peak frequency, reduced alpha activity, and localized increases in broadband gamma power. We speculate in females with FXS, relatively intact thalamocortical function leads to increase alpha power to compensate for elevated gamma ‘noise’ (based on shared genetic vulnerability). This speculation is supported by the higher SA ratio in females with FXS and relatively spared auditory attention performance.

The SA ratio was our attempt to capture the relationship between neural synchronization and background asynchronous activity. This ratio is the degree of gamma phase synchronization (40 Hz ITPC) weighted by the amount of gamma “noise” (gamma1 STP). We hypothesized that a higher SA ratio would represent a higher signal-to-noise of functional neural activity. We found significantly lower SA ratios in males with FXS and across all FXS subjects a high degree of association with increased SA ratio and increased auditory attention performance. We speculate in FXS, engagement of a cortical circuit (i.e., input sound) leads to increased activity (i.e., gamma STP) but does not lead to the expected phase reset of gamma oscillations (i.e., 40 Hz ITPC). Though the implementation of SA ratio will vary by task, the current data demonstrates the clinical utility of a conceptual SNR measures as a functional alternative in humans where excitatory/inhibitory metrics cannot be easily measured. Future development of the measure may include assessing responsiveness to intervention or study between molecular subtypes of FXS.

### Limitations

In our analysis, we reported absolute power which can vary more between subjects. This simplifies interpretation since power ratios in FXS strongly diverge from controls across multiple bands. The MNE model generates a current dipole, in the units of A-m, for each vertex which is a spatially filtered and greatly attenuated compared to the original scalp amplitude. We recently showed that relative and absolute resting source power has comparable directional relationship between FXS and controls^61^. ITPC measures were also used and are standardized within each frequency. We used a standardized, averaged anatomy (ICBM152) to compute the electric lead field as no individual anatomies were available. Previous efforts for large scale magnetic resonance imaging in FXS shows a bias towards females or males with higher cognitive function, thus introducing potential bias. If the results are interpreted with care, EEG source modeling can be justified without individual anatomical information^15, 62^. We opted to study cortical regions which considered multiple, localized atlas nodes as replicates to reduce spatial precision, but more accurately estimate brain activity of larger areas.

## Conclusion

The use of source localization to trace findings to distinct neural regions can aid in the development of biomarkers and further advance preclinical studies. The ratio of synchronous and asynchronous gamma activity (SA ratio) may further represent a clinically composite biomarker of the efficiency of auditory processing in FXS. Findings from this study have implications for back translation and relevance for signal-to-noise conceptual frameworks in understanding neural activity in neurodevelopmental disorders.

## Data Availability

All data produced in the present study are available upon reasonable request to the authors

---

**Supplemental Figure 1:**
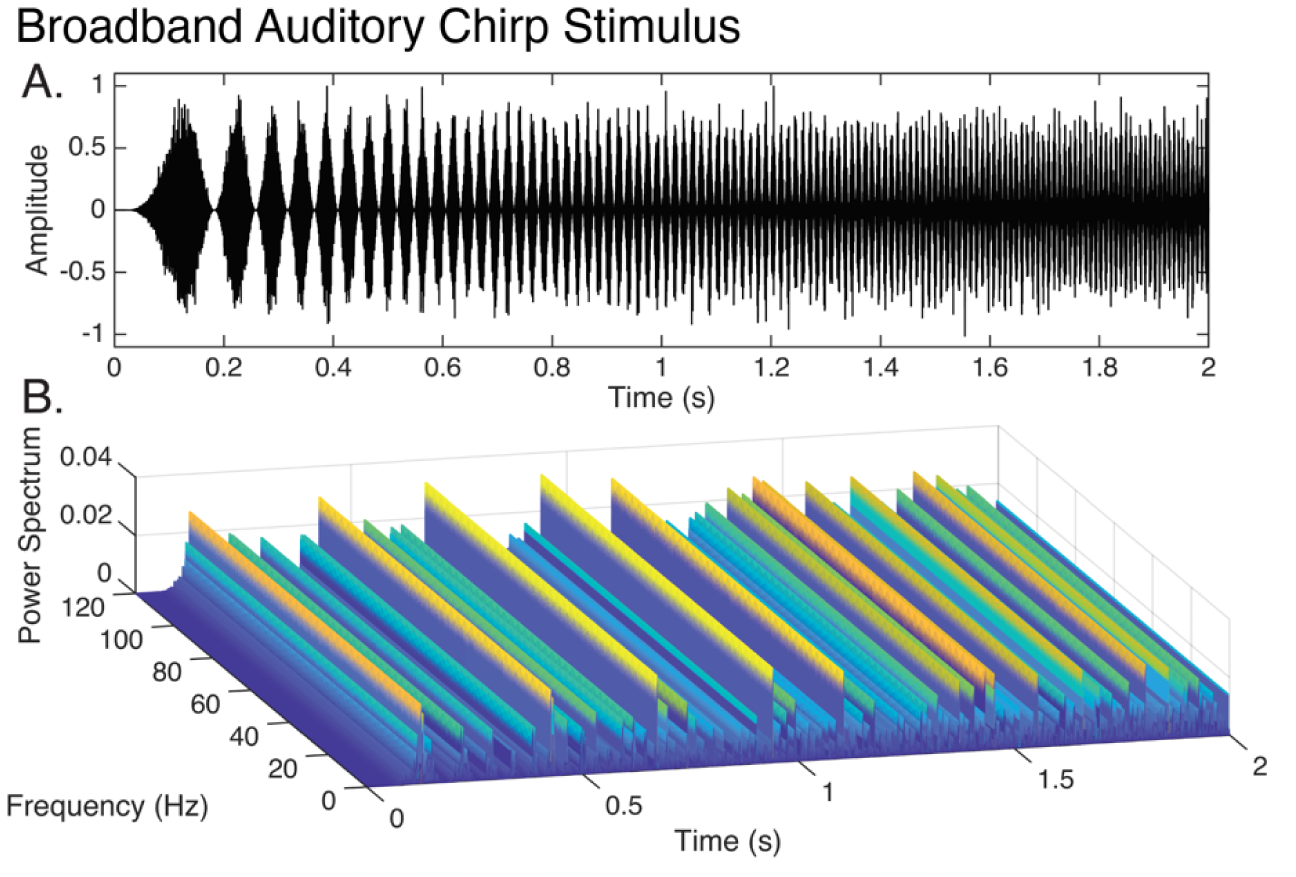
Broadband auditory chirp stimulus. A. Plot of amplitude-modulated pink noise carrier waveform. Frequency of modulation increases from 0 to 120 Hz over 2 seconds. B. Waterfall plot depicting power spectrum of broadband chirp stimulus.

**Supplemental Figure 2:**
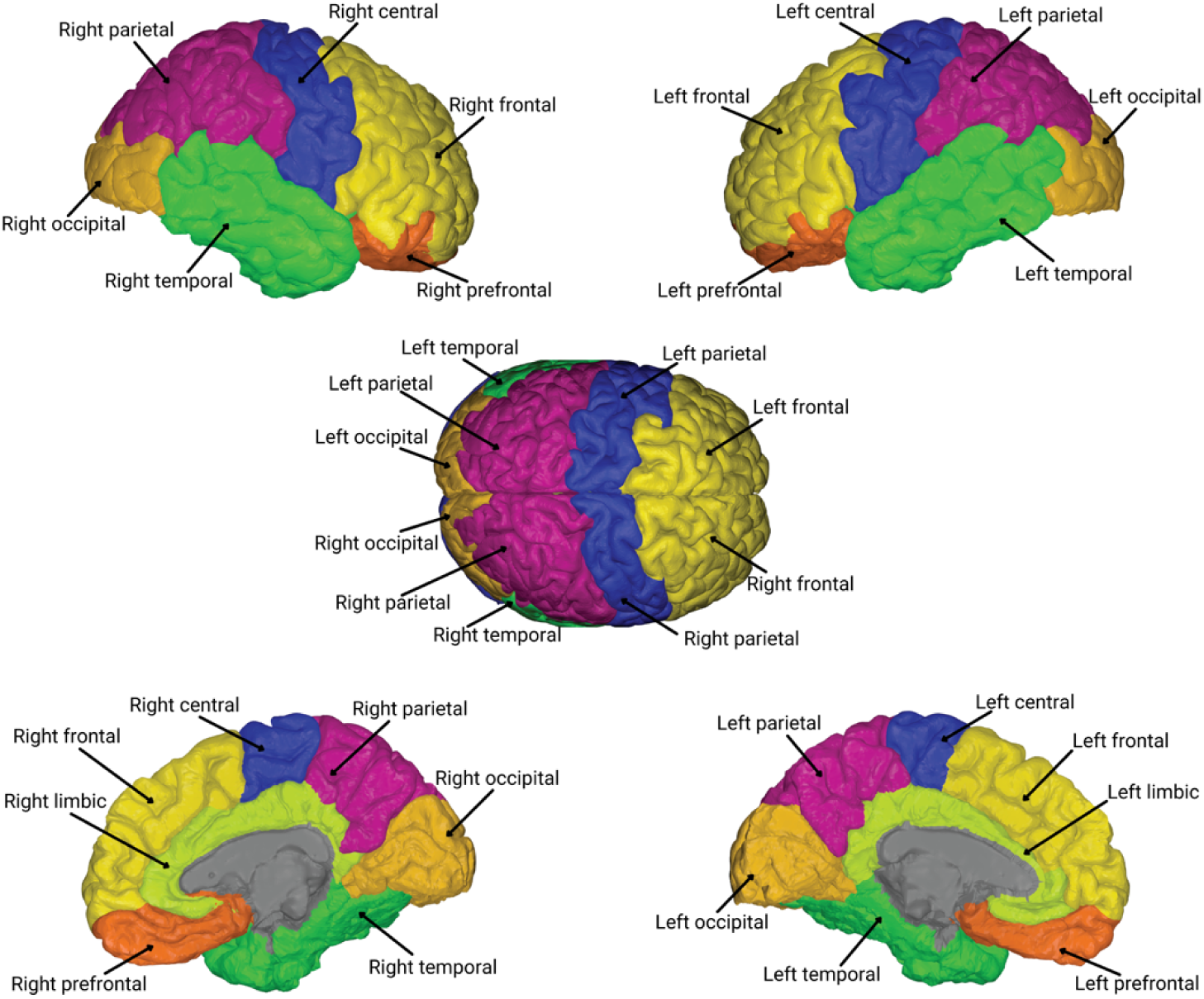
Atlas visualization of Desikan-Killiany based cortical regions.

**Supplemental Table 3:**
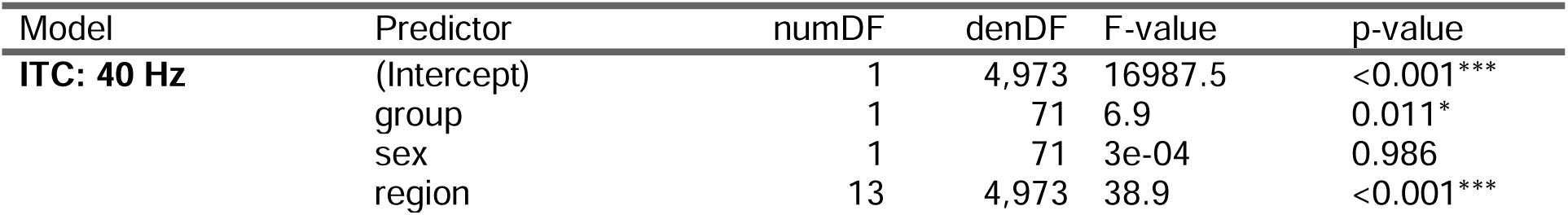
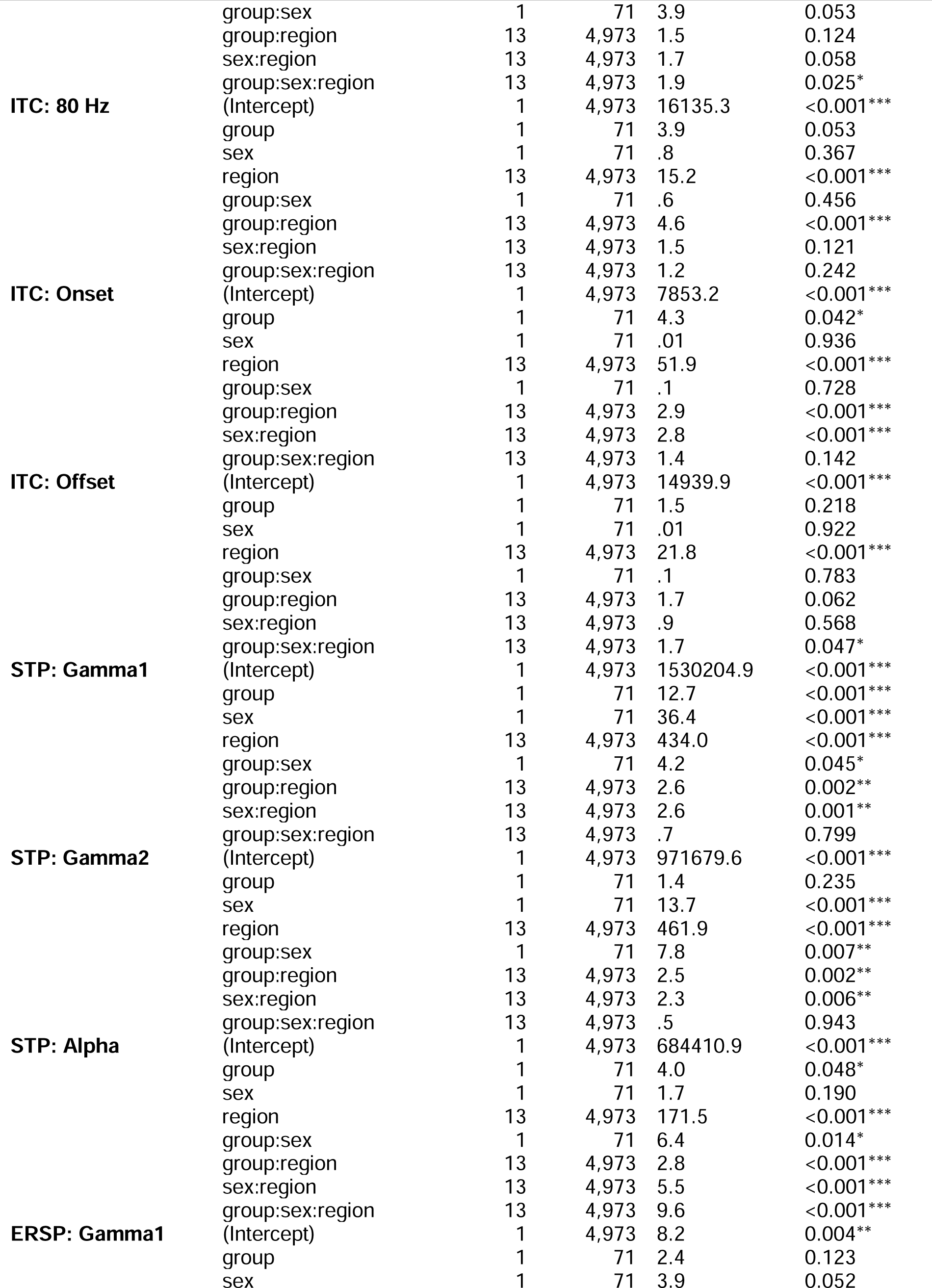
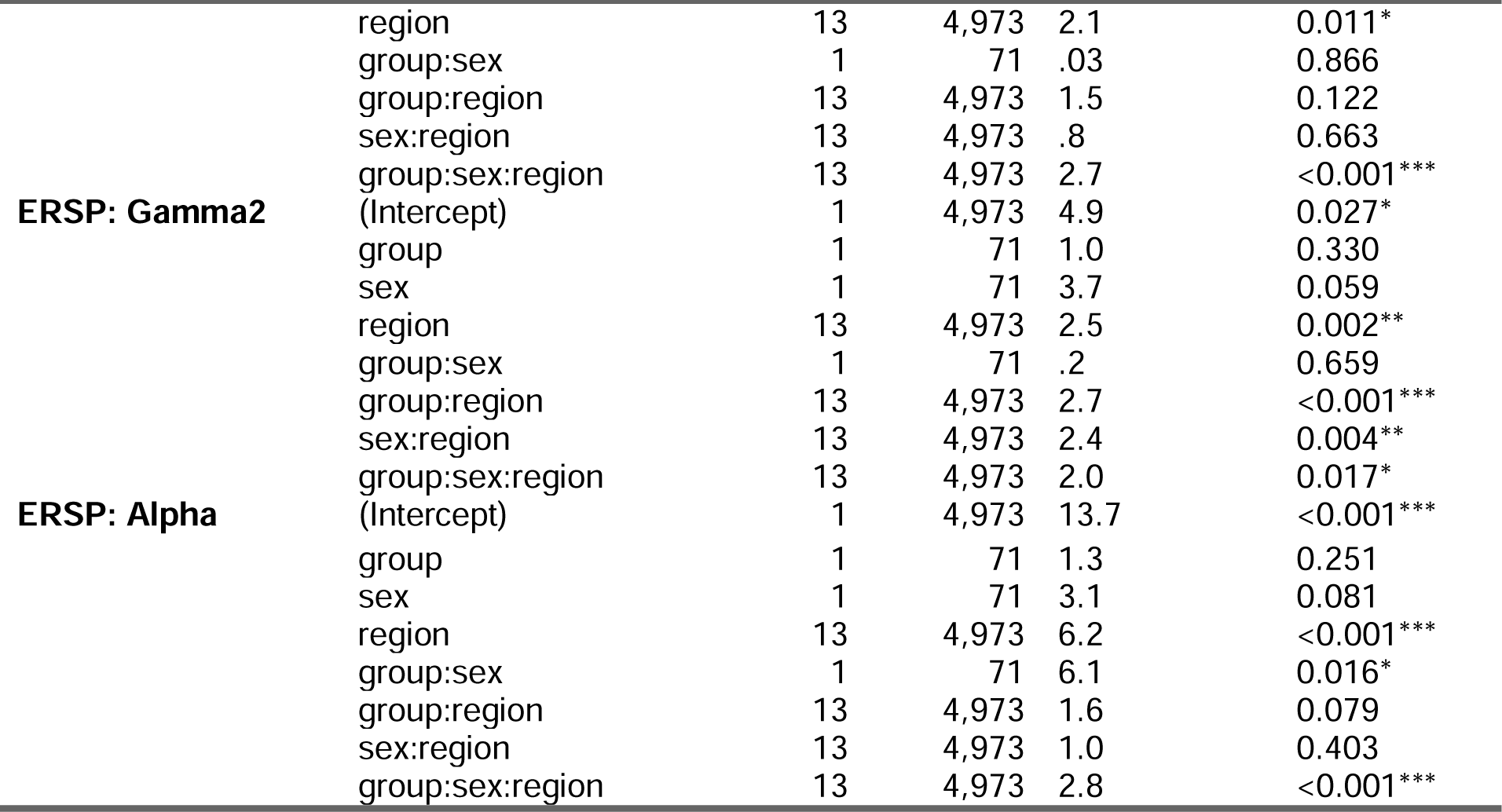
Results of Linear Mixed Effect models of key EEG variables.

